# Performance of the TaqMan™ COVID-19 Pooling Kit for detection of SARS-CoV-2 in Asymptomatic and Symptomatic populations at an Institution of Higher Education

**DOI:** 10.1101/2021.05.20.21257523

**Authors:** Troy Ganz, Sarah Sanderson, Connor Baush, Melanie Mejia, Manoj Gandhi, Jared Auclair

## Abstract

Clinical evidence for asymptomatic cases of coronavirus disease (COVID-19) has reinforced the significance of effective surveillance testing programs. Quantitative reverse transcriptase polymerase chain reaction (RT-qPCR) assays are considered the ‘gold standard’ for detection of severe acute respiratory syndrome coronavirus 2 (SARS-CoV-2) RNA. However, the labor and resource requirements can be prohibitive with respect to large testing volumes associated with the pandemic. Pooled testing algorithms may serve to increase testing capacity with more efficient resource utilization. Due to the lack of carefully curated cohorts, there is limited evidence for the applicability of RT-PCR pooling in asymptomatic COVID-19 cases. In this study, we compared the analytical sensitivity of the TaqMan™ SARS-CoV-2 Pooling Assay to detect one positive sample in a pool of five anterior nare swabs in symptomatic and asymptomatic cohorts at an institute of higher education. Positive pools were deconvoluted and each individual sample was retested using the TaqPath™ COVID-19 Combo Kit. Both assays target the open reading frame (ORF) 1ab, nucleocapsid (N), and spike (S) gene of the strain that originated in Wuhan, Hubei, China. Qualitative results demonstrated absolute agreement between pooled and deconvoluted samples in both cohorts. Independent t-test performed on C_t_ shifts confirmed an insignificant difference between cohorts with p-values of 0.306 (Orf1ab), 0.147 (N), and 0.052 (S). All negative pools were correctly reported as negative. Thus, pooled PCR testing up to five samples is a valid method for surveillance testing of students and staff in a university setting, especially when the prevalence is expected to be low.

## Introduction

Efforts to mitigate the reproductive number (R_0_) for the coronavirus disease 2019 (COVID-19) pandemic, an emergent respiratory infection caused by severe acute respiratory syndrome coronavirus 2 (SARS-CoV-2), has been undermined by the potential for sustained transmission through pre-symptomatic, paucisymptomatic (subclinical), and asymptomatic carriers [1]. Growing evidence from immunological case studies and bioinformatic investigations have substantiated clinical diagnostic testing that have reported insignificant variance in results among patients that differ in symptomology. Asymptomatic patients have reportedly developed ground-glass opacities in lung tissue and significant adaptive immune (IgG and IgM) responses, however cell-mediated immune response from asymptomatic cases have shown to be less significant compared to symptomatic cases [2,3]. A recent investigation noted a correlation between globally reported asymptomatic cases and a single nucleotide polymorphism (SNP) in SARS-CoV-2 RNA at position 11083G>T translating to a phenylalanine mutation (L37F) on non-structural protein 6 (NSP6) [4]. While a less severe immune response may be elicited in asymptomatic cases, the replication mechanisms in these individuals remain intact, and this necessitates effective risk management measures [5].

The analytical sensitivity of RT-qPCR has propelled its utility as the ‘gold standard’ to accurately detect SARS-CoV-2 RNA from upper respiratory specimens (e.g., nasopharyngeal, anterior nare, and saliva); however, the high-complexity format has merged with demands of a pandemic to generate severe shortages in materials and technical personnel. A resurgence of interest in group testing, or sample pooling, whereby the ratio of specimens to result is greater than one, has led investigators to optimize protocols that would allow an increase in testing capacity without requiring additional resources [6]. Caveats include a linear loss of sensitivity with sample dilution, and, in hierarchical systems, disease prevalence may determine efficiency as positive pools are deconvoluted to retest each patient individually [7]. Nevertheless, sample pooling stands to provide an accurate and affordable method for general population testing.

Since gaining USFDA emergency use authorization (EUA) for the TaqPath™ COVID-19 Combo Kit on March 24, 2020, Thermo Fisher Scientific, Inc. has developed a specialized RT-qPCR testing kit with enhanced sensitivity for the purposes of pooling up to five samples known as the TaqMan™ SARS-CoV-2 Pooling Assay. In the following study, we compared the sensitivity of the TaqMan™ assay to detect one positive sample in a pool of five from 30 symptomatic and 30 asymptomatic individuals that were previously characterized as positive using the TaqPath™ COVID-19 Combo Kit; 10 negative pools of five were included as a control group.

## Methods

### Sample Collection, Storage, and Preanalytical Processing

The study was conducted at the Northeastern University COVID-19 response laboratory, the Life Science Testing Center (LSTC; Burlington, MA, USA), which performs population testing for university affiliates. Students on campus are required to test every three days while staff and faculty on campus test twice a week. Affiliates attending campus are required to complete a daily wellness tracker, or a two-question survey, that identifies whether they have experienced COVID-19 related symptoms or have been in close contact with a positive COVID-19 case in the past 24 hours. Depending on the survey results, they are directed to get tested at either the non-symptomatic site (Cabot Center), where observed self-collection is performed, or the symptomatic site (Marino Center), where collection is performed by a healthcare professional. Temperature checks and general symptom screening were performed by trained personnel on the subjects at both sites to determine presence or absence of symptoms. Anterior nasal swabs (polyester) are collected in 3 mL BD Vacutainer® (without additive) tubes and transported dry to the LSTC facility. Upon logging into the laboratory information system (LIMS), 3 mL of viral transport media (VTM; Redoxica, Little Rock, AR, USA) is added and the sample is shaken at 1000 rpm for 5 minutes. Processed specimens are stable for <72 hours at 2-8°C. Positive samples are catalogued, split into three aliquots, and stored indefinitely at -80°C. Depending on the site at which the samples are collected (Cabot or Marino), this monitoring approach allows for curated cohorts of symptomatic or asymptomatic cases.

### TaqMan™ SARS-CoV-2 Pooling Assay: Pooled Specimen RNA Extraction and RT-qPCR Setup

Pools consisting of five samples were constructed from one positive sample and four negative samples. Positive samples were selected at random from -80°C storage and thawed at 2-8°C. RNA extraction was performed using 400 µL of combined sample volume using the MagMAX™ Viral/Pathogen II (MVP II) Nucleic Acid Isolation Kit (Thermo Fisher Scientific, Waltham, MA, USA) by combining 10 µL of Proteinase K, 80 µL of each sample (400 µl pooled volume), 550 µL of lysis buffer with RNA binding beads, and 10 µL of MS2 phage extraction control to a single well of an Agilent 1 mL 96-well plate. The plate was vortexed 2 minutes at 1050 rpm, incubated 5 minutes at 65°C, vortexed for 5 minutes at 1050 rpm, and incubated at room temperature on a magnetic stand. After waste aspiration, the beads underwent three cycles of resuspension/aspiration in 1 mL wash buffer, 1 mL 80% ethanol, and 50 µL elution buffer, respectively. RT-qPCR was performed using 17.5 µL of eluant added to 7.5 µL of reaction mix that contained 6.25 µL TaqPath™ 1-Step RT-qPCR MM, CG (4X) and 1.25 µL TaqMan™ SARS-CoV-2 Pooling Multiplex Assay.

### TaqPath™ COVID-19 Combo Kit: Deconvoluted Specimen RNA Extraction and RT-qPCR Setup

Pools that tested positive were deconvoluted and each of the five component specimens were repeated using the TaqPath™ COVID-19 Combo Kit currently employed at the LSTC. Briefly, RNA extraction was performed using the MVPII kit with 5 µL of Proteinase K, 200 µL sample, 275 µL lysis buffer with RNA binding beads, and 5 µL MS2 phage extraction control. Wash procedure was performed using an Agilent Bravo liquid handler (Agilent Technologies, Santa Clara, CA, USA) whereby, after initial shaking 2 minutes at 1050 rpm and 5 minutes incubation at 65°C, the beads underwent three cycles of resuspension/aspiration in 165 µL wash buffer, 165 µL 80% ethanol, and 50 µL elution buffer, respectively. RT-qPCR was performed using 10 µL of eluate added to 15 µL of reaction mix that contained 6.25 µL TaqPath™ 1-Step Multiplex Master Mix (No ROX™) (4x), 1.25 µL COVID-19 Real Time PCR Assay Multiplex, and 7.5 µL Nuclease-free water.

### RT-qPCR Thermal Profile and Results Management

Both, the TaqMan™ and TaqPath™, methods contain primers and probes that target the SARS-CoV-2 open reading frame (ORF) 1ab, nucleocapsid (N), and spike (S) genes as well as the MS2 phage extraction control (reporter dyes FAM, VIC, ABY, and JUN, respectively). Thermal profile parameters for both assays involved a 2-minute UNG incubation cycle at 25°C, a 10-minute reverse transcriptase incubation cycle at 53°C, a 2-minute activation cycle at 95°C, and 40 cycles of a 3-second denaturation at 95°C followed by 30-second anneal/extension cycle at 60°C on Applied Biosystems™ 7500 Fast Dx Real-Time PCR Instrument (Thermo Fisher Scientific, Waltham, MA, USA). Resulting threshold cycle (C_t_) values were extracted using COVID-19 Interpretive Software and analyzed using Microsoft Excel® and GraphPad Prism version 9 for Windows (GraphPad Software, San Diego, CA, USA). Qualitative endpoints set to determine target presence were C_t_≤37 for SARS-CoV-2 genes and C_t_≤32 for MS2 control; ≥2/3 SARS-CoV-2 genes designated a positive result and 1/3 SARS-CoV-2 genes designated an indeterminate result.

## Results

Average monthly prevalence estimates ranging August 2020 to April 2021 for the Cabot Center (non-symptomatic) was 0.159% (CI.95 0.086-0.234%) and Marino Center (symptomatic) was 3.33% (CI.95 2.05-4.61%). Thirty samples from each of the two sites were used to generate 60 five-sample pools with 240 negative samples. Each of the 60 five-sample pools tested positive using the TaqMan™ pooling method and statistical analysis was performed using the deconvoluted result. Average C_t_ values for the pooled asymptomatic samples were 22.17 (CI.95 20.18-24.16), 22.66 (CI.95 20.76-24.57), and 23.15 (CI.95 21.13-25.17) and the deconvoluted asymptomatic samples were 20.64 (CI.95 18.59-22.68), 21.30 (CI.95 19.38-23.22), and 20.81 (CI.95 18.92-22.69) for the ORF1ab, N, and S genes, respectively (Figure1: a, b, c). Average C_t_ values for the pooled symptomatic samples were 22.47 (CI.95 20.31-24.63), 23.31 (CI.95 21.19-25.44), and 24.01 (CI.95 21.15-26.87) and the deconvoluted symptomatic samples were 20.65 (CI.95 18.53-22.77), 21.70 (CI.95 19.60-23.80), and 21.17 (CI.95 18.63-23.71) for the ORF1ab, N, and S genes, respectively (Figure1: d, e, f). Dependent t-test analysis comparing pooled to deconvoluted data sets indicated a statistically significant difference with p-values of <0.001 for all comparisons, however this is reflective of the expected effect on C_t_ value due to dilution. Average C_t_ shifts of 1.54±1.07 (ORF1ab), 1.37±0.77 (N), and 2.34±1.68 (S) for the asymptomatic cohort and 1.82±1.01 (ORF1ab), 1.62±0.48 (N), and 3.10±1.37 (S) for the symptomatic cohort was observed. Independent t-test did not show a statistically significant C_t_ shift between the asymptomatic and symptomatic cohorts with p-values of 0.306, 0.147, and 0.052 for ORF1ab, N, and S genes, respectively (Figure 1g). Each negative pool reported negative and were not deconvoluted.

**Figure 1:**
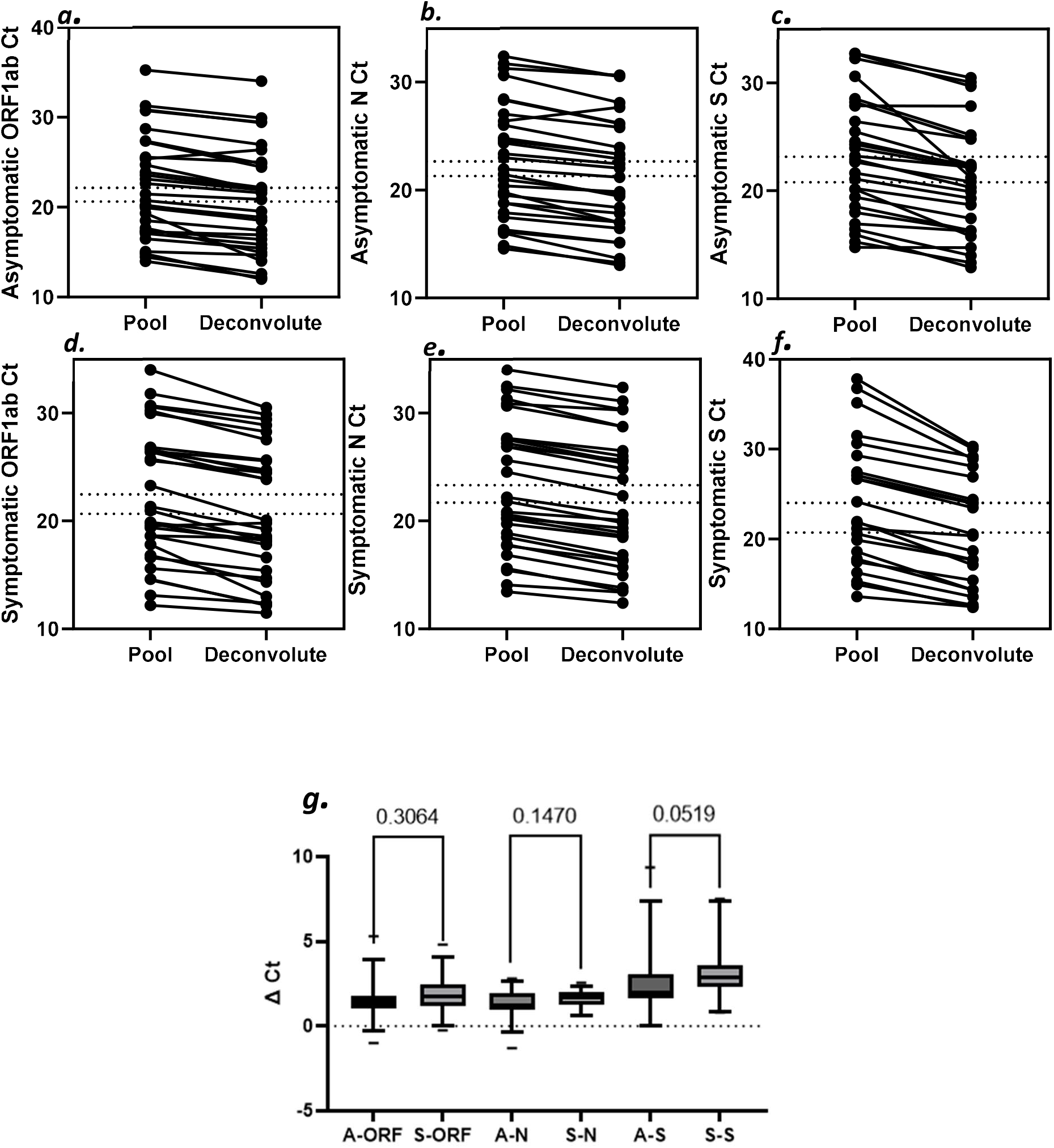
Pairwise comparisons of pooled and deconvoluted samples for each gene target (ORF1ab, N, and S) from Asymptomatic (*a,b,c*) and Symptomatic cohorts (*d,e,f*); means indicated by dashed intersects. Box-and-whisker plot describing C_t_ shift for each gene from each cohort; bars are CI.95; ranges marked; p-values annotated (*g*).

## Discussion

This study sought to confirm SARS-CoV-2 infections that cause mild immune responses proliferate to similar extents as those that elicit severe cases of COVID-19, which enables equal probability of detection in RT-qPCR assay. Furthermore, the TaqMan™ multiplex assay identified each positive sample in pools of five with minimal loss of sensitivity. Prevalence of SARS-CoV-2 variants, apparent by S gene target loss, has increased by an order of magnitude since November 2020 at the LSTC and cases represented 7% of asymptomatic and 20% of symptomatic cases in the sample size for this study [8]. It is important to note, significant dilution effects observed in all genes was a result of interpreting one positive per pool of five and may indicate a limitation of the experimental design due to probability of more than one positive per pool in practice [9]. Thus, the practical significance observed by the average C_t_ shifts and negligible impact to qualitative agreement supports retention of RT-qPCR sensitivity at a minimum.

This strategy of increased extraction and elution volumes for group tests reported absolute qualitative agreement to the deconvoluted counterpart, including a 1/3 gene positive case, and produced a more conserved C_t_ shift [7,9]. Since symptomatic subjects tend to have a significantly higher prevalence of COVID-19 in a pandemic setting, from a resource efficiency standpoint, it is advisable to consider pooling samples when testing subjects who are non-symptomatic such as in vaccinated populations, where the prevalence rates are expected to be significantly lower.

## Data Availability

The authors confirm that the data supporting the findings of this study are available within the article.

## Ethical Statement

This work was performed under Northeastern University Office of Human Subject Research Protection (HSRP) IRB approval.

## Acknowledgements

The authors would like to acknowledge the efforts of the LSTC technical staff, including M.K., R.M. who made contributions to this work.

